# Automatic Quantification of Ki-67 Labeling Index in Pediatric Brain Tumors Using Qupath

**DOI:** 10.1101/2025.05.09.25327292

**Authors:** Christoforos Spyretos, Juan Manuel Pardo Ladino, Hakon Blomstrand, Per Nyman, Oscar Snödahl, Alia Shamikh, Nils O. Elander, Neda Haj-Hosseini

## Abstract

The quantification of the Ki-67 labeling index (LI) is critical for assessing tumor proliferation and prognosis in tumors, yet manual scoring remains a common practice. This study presents an automated workflow for Ki-67 scoring in whole slide images (WSIs) using an Apache Groovy code script for QuPath, complemented by a Python-based post-processing script, providing cell density maps and summary tables. The tissue and cell segmentation are performed using StarDist, a deep learning model, and adaptive thresholding to classify Ki-67 positive and negative nuclei. The pipeline was applied to a cohort of 632 pediatric brain tumor cases with 734 Ki-67-stained WSIs from the Children’s Brain Tumor Network. Medulloblastoma showed the highest Ki-67 LI (median: 19.84), followed by atypical teratoid rhabdoid tumor (median: 19.36). Moderate values were observed in brainstem glioma-diffuse intrinsic pontine glioma (median: 11.50), high-grade glioma (grades 3 & 4) (median: 9.50), and ependymoma (median: 5.88). Lower indices were found in meningioma (median: 1.84), while the lowest were seen in low-grade glioma (grades 1 & 2) (median: 0.85), dysembryoplastic neuroepithelial tumor (median: 0.63), and ganglioglioma (median: 0.50). The results aligned with the consensus of the oncology, demonstrating a significant correlation in Ki-67 LI across most of the tumor families/types, with high malignancy tumors showing the highest proliferation indices and lower malignancy tumors exhibiting lower Ki-67 LI. The automated approach facilitates the assessment of large amounts of Ki-67 WSIs in research settings.

## 1 Introduction

In histopathology, hematoxylin and eosin (H&E) staining is the gold standard for morphological assessment of various tissue types, including cancer. In addition to H&E, pathologists often use antibody-dependent immunohistochemical (IHC) protocols to identify specific molecular alterations, assess expression levels and distribution of various proteins, and determine molecular subgroups of cancers [1]. One such IHC marker is the Antigen Kiel 67 protein (Ki-67), which is known for distinguishing between proliferating and non-proliferating cells. Ki-67 is expressed by nuclear staining and detected in every active phase of the cell cycle, whereas it is absent during the temporary phase. In this regard, the ratio of the number of positive cells to the total number of cells, known as the Ki-67 labeling index (LI), is a key component in assessing the character of diverse tumors [2, 3]. Ki-67 can specifically be valuable in low-resource settings where the molecular examination, which provides a more accurate diagnosis, is inaccessible.

Given Ki-67’s extensive use in oncological management, it has also been widely studied in adult, pediatric, and adolescent central nervous system (CNS) tumors for providing information on malignant potential and prognosis. More precisely, an increased Ki-67 LI is associated with higher malignancy in gliomas and is used to differentiate between low-grade and high-grade gliomas. In addition, it is a valuable prognostic factor for estimating tumor progression and survival [4, 5, 6]. Regarding glioblastoma patients, the relationship between Ki-67 LI and overall survival, and its role in distinguishing subtypes remains unclear [5, 6, 7]. In the pediatric population, a correlation has also been observed between the ependymoma grades and Ki-67 LI, and between the prognosis of ependymoma and medulloblastoma with Ki-67 LI [8, 9].

The percentage of the Ki-67 LI is reported to correlate with the proliferation rate and aggressiveness of the cancer and, consequently, the survival prognosis [2]. Despite a straightforward measure of the proliferation index provided by Ki-67 LI, there is an absence of standardized method for counting stained nuclei, and selecting a representative and adequately sized area (hot spot) in a possibly heterogeneously proliferating tumor. Therefore its clinical utility has been constrained, resulting in poor interobserver reproducibility and high rating variability of the LI [10, 11, 12]. While several studies have demonstrated the benefits of digital image analysis for assessing the Ki-67 LI, there are considerable discrepancies about how to count the cells [5, 13]. The most reported drawback is the risk of counting non-neoplastic cells, such as lymphocytes, microglia, and macrophages, as well as other brown-pigmented signals, such as hemosiderin and hematoidin. Thus, non-neoplastic cells might contribute to the Ki-67 LI and influence the assessment and interpretation.

These errors can be solved by having pathologists manually annotate regions of interest, or through IHC double-staining with both Ki-67 and a tumor cell-specific marker, but both approaches are labor-intensive and cost-driving. Standardizing the concept of tumor areas with Ki-67-stained nuclei is an essential parameter in calculating the LI. Deep learning methods have been suggested as more powerful tools that can be used for automatic cell segmentation [14, 15] and for potentially solving such issues [16, 17]. However, integrating of these models into user-friendly interfaces or widely used pathology software is essential. Among the few non-commercial alternatives, DeepLIIF performs automated cell segmentation and classification on IHC images [18], with most commercial platforms not being suitable for large-scale data analysis, leading to limited accessibility.

This study aimed to develop scripts for automatic scoring of Ki-67 LI in a commonly used software that can analyze relatively large amounts of images without user intervention and evaluate its performance. In this regard, Apache Groovy (Java-based syntax) code script for QuPath [19, 20], an open-access software, was developed to automate cell counting across multiple pediatric brain tumor families/types using WSIs.

## 2 Materials and Methods

### 2.1 Data

In this study, the open-access dataset was obtained upon approval from the Children’s Brain Tumor Network (CBTN) in 2023 [21, 22]. The dataset included various stained WSIs, with H&E being the most common and Ki-67 being one of the most representative IHC staining protocols. Furthermore, the obtained version of the CBTN dataset is based on pre-2021 WHO guidelines, in which some of the brain tumor classifications are no longer used in clinical practice [23]. Therefore, in accordance with clinicians, subjects with outdated classifications were excluded, and the classifications listed in the 2021 WHO guidelines were used to conduct the experiments. The study analyzes the tumor families of low-grade glioma (grades 1, 2) (LGG) and high-grade glioma (grades 3, 4) (HGG), and the cancer types of medulloblastoma (MB), ependymoma (EP), ganglioglioma (GG), meningioma (MEN), atypical teratoid rhabdoid tumor (ATRT), dysembryoplastic neuroepithelial tumor (DNET) and brainstem glioma-diffuse intrinsic pontine glioma (DIPG). Additionally, 139 slides containing control tissue on the left side were cropped to analyze only the right side, an example of a control tissue image is depicted in Figure 3. In total, 629 subjects were included in the analysis; however, the diagnosis of three subjects changed over time and therefore were treated as independent cases, resulting in 632 cases (male/female: 339/293; mean age in years: 10.43 ± 6.55 years) and 734 Ki-67 WSIs, with some cases having more than one slide. Additionally, tumor descriptor information for each case was extracted from the corresponding magnetic resonance imaging (MRI) dataset, with 413 cases classified as initial CNS tumors, 92 as progressive, 40 as recurrent, and 12 as second malignancy, while 104 tumors did not have any descriptor. Among the cases, 29 had two tumor descriptors and one case had three tumors descriptors. It should be noted that there is no a one-to-one correspondence in the rest of the cases between the initial CNS and later tumors descriptors.

**Figure 1.**
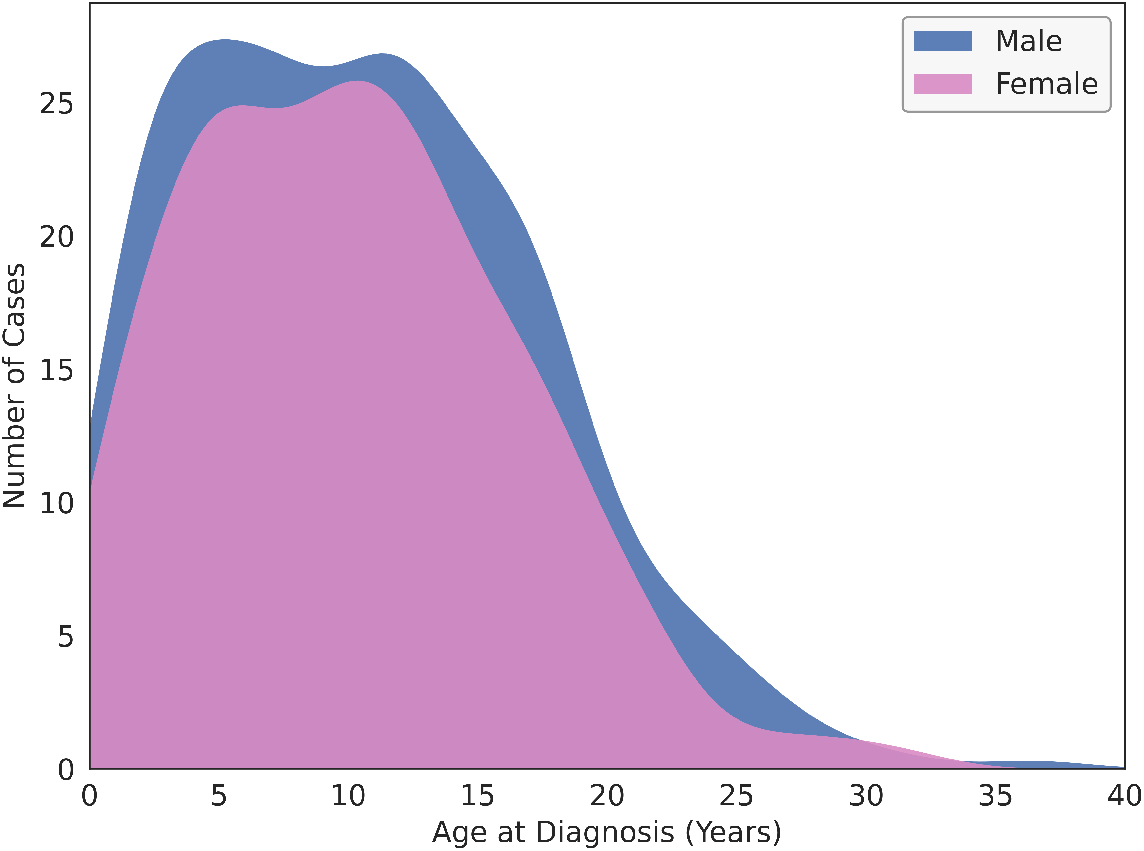
Age distribution of male and female cases (total: 632 cases, males: 339, females: 293) with a mean age of 10.43 ± 6.55 years.

**Figure 2.**
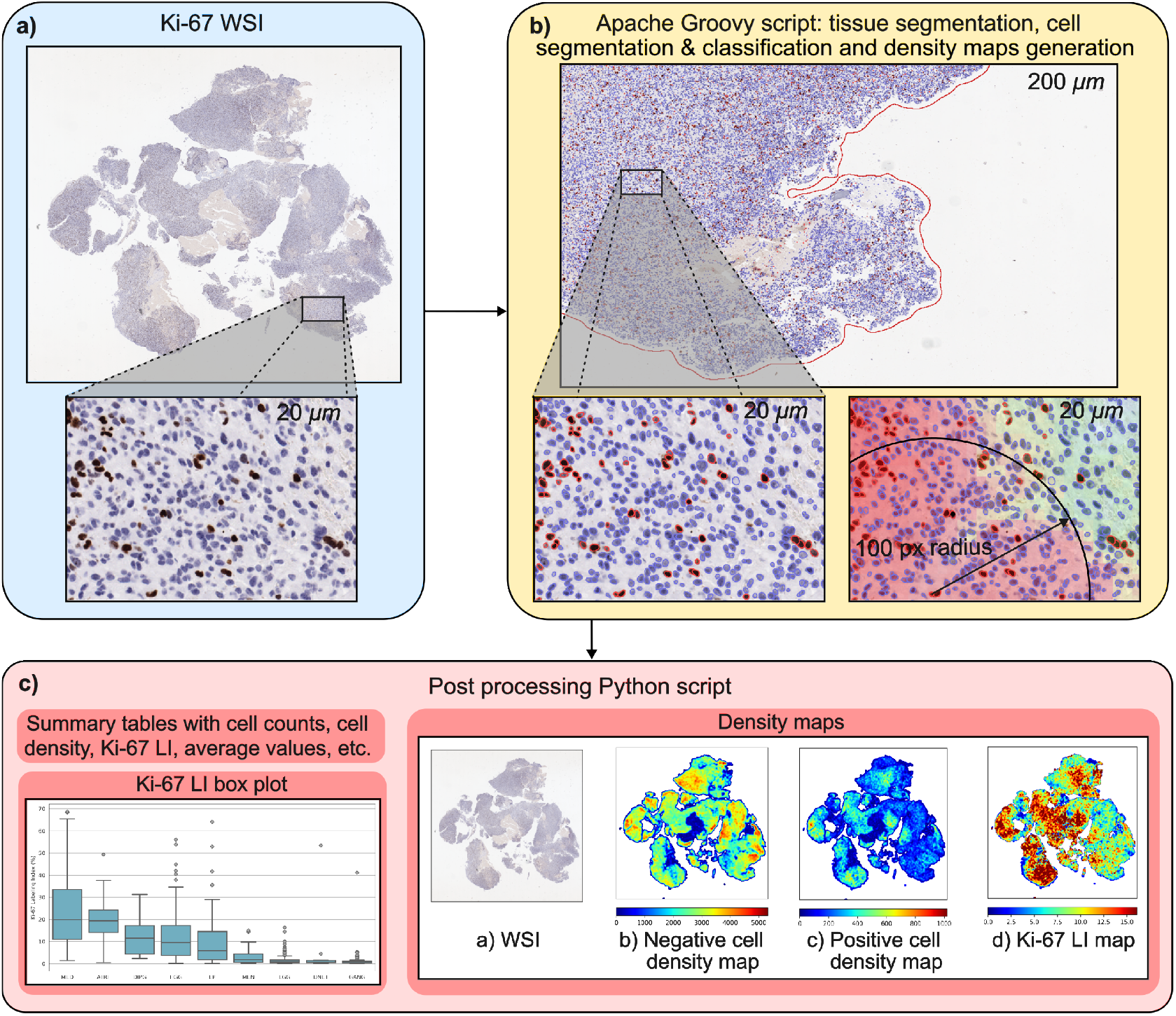
Overview of the workflow. **a)** Ki-67 WSIs are imported into QuPath. Representative Ki-67 WSIs with 20 micrometers (*µm*) zoomed-in region from a subject diagnosed with HGG. Ki-67 negative nuclei are stained blue, while positive nuclei are stained brown. **b)** Tissue segmentation, and cell segmentation and classification are performed by the Apache Groovy script. Image metadata and cell density maps are extracted using the classified nuclei with a search radius of 100 pixels (50*µm*) and stored. **c)** Density maps are processed, and summary graphs and tables are produced by Python script.

**Figure 3.**
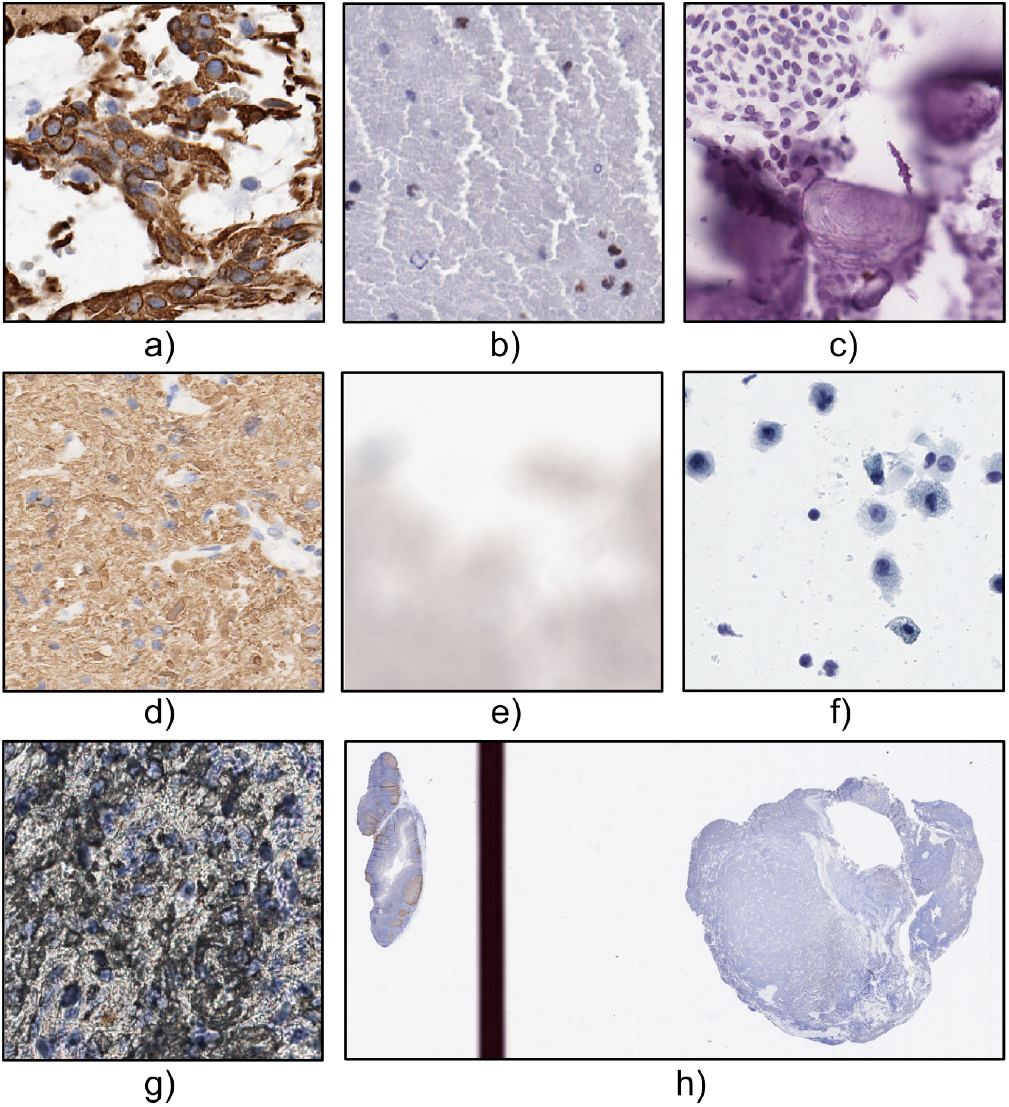
Example of cases where cell classification was erroneous at x20 magnification. **a)** Stained cytoplasm and axons, **b)** clumped erythrocytes, **c)** bone fragments, **d)** background staining, **e)** out-of-focus WSI, **f)** light exposure tissue, and **g)** artifact during tissue preparation. **h)** Example of a WSI with a control tissue sample on the left part. Control tissue samples serve as a reference point when tissue samples are examined.

Table 1 summarizes the final number of subjects and WSIs, and Figure 1 depicts the age distribution.

**Table 1.** Number of subjects and Ki-67 WSIs for each tumor family/type included in the study.

| Tumor Family/Type | Ki-67 |  | Ki-67 with Tumor Descriptor |  |
| --- | --- | --- | --- | --- |
|  | No. of Subjects<br>(Male/Female) | No. of WSIs | No. of Subjects<br>(Male/Female) | No. of WSIs |
| Low-grade glioma<br>(grades 1, 2) (LGG) | 272<br>(147/125) | 309 | 272<br>(147/125) | 274 |
| High-grade glioma<br>(grades 3, 4) (HGG) | 88<br>(48/40) | 103 | 76<br>(43/33) | 90 |
| Medulloblastoma (MB) | 81<br>(46/35) | 86 | 75<br>(42/32) | 79 |
| Ependymoma (EP) | 71<br>(45/26) | 89 | 59<br>(37/22) | 75 |
| Ganglioglioma (GG) | 51<br>(28/23) | 65 | 41<br>(23/18) | 54 |
| Meningioma (MEN) | 24<br>(13/11) | 34 | 5<br>(4/1) | 5 |
| Atypical teratoid rhabdoid tumor (ATRT) | 20<br>(13/7) | 21 | 18<br>(12/6) | 19 |
| Dysembryoplastic neuroepithelial<br>tumor (DENT) | 14<br>(11/3) | 15 | 1<br>(1/-) | 1 |
| Brainstem glioma-diffuse intrinsic<br>pontine glioma (DIPG) | 11<br>(7/4) | 12 | 9<br>(7/2) | 10 |
| Total | 632<br>(339/293) | 734 | 528<br>(276/252) | 607 |

### 2.2 Analysis Scripts

The analysis of the images was performed using an Apache Groovy script in QuPath, and a Python script was used for post-processing, which are user-friendly, requiring minimal coding expertise. The Groovy script is designed to perform an initial tissue segmentation using a predefined pixel classifier to eliminate artifacts, followed by refinement of stain vector estimation. In the next step, a threshold-based classifier is applied to achieve refined tissue segmentation based on the updated stain parameters. Subsequently, cell segmentation is performed, after which cells are classified as positive or negative, while image metadata is stored, and cell density maps are generated. The Python codes further process the density maps by normalizing and visualizing them, and producing summary graphs and tables of the Ki-67 LI. Users can run the Python script via the terminal by specifying a set of defined arguments. The complete analysis workflow is illustrated in Figure 2.

#### 2.2.1 Apache Groovy Script Functions

##### Preliminary Tissue Segmentation

Each WSI was initially set to the Brightfield_H_DAB image type, with defined generic color deconvolution stain vectors. DAB stands for diaminobenzidine, an organic compound used in IHC stains. In addition, a predefined pixel classifier was selected in QuPath, named Base_Classifier, which is applied to eliminate artifacts such as dark areas and pen marks. The values for the minArea and minHoleArea were selected through trial and error to achieve an optimal balance between minimizing noise and capturing tissue regions. The default parameter settings for this preliminary tissue segmentation are outlined in Table 2.

**Table 2.** Default and adapted parameter settings for the pixel classifiers of the preliminary and threshold-based tissue segmentations.

| Parameter | Preliminary Segmentation | Threshold-Based Segmentation |
| --- | --- | --- |
| Pixel classifier type (default) | OpenCVPixelClassifier | OpenCVPixelClassifier |
| Input padding | 0 | 0 |
| Input resolution | Pixel width: 4.0016 $\mu m$<br>Pixel height: 4.0016 $\mu m$<br>Z spacing: 1.0 z-slice | Pixel width: 4.0016 $\mu m$<br>Pixel height: 4.0016 $\mu m$<br>Z spacing: 1.0 z-slice |
| Input dimensions (default) | Input width: 512 pixels<br>Input height: 512 pixels<br>Input number of channels: 3 (RGB values) | Input width: 512 pixels<br>Input height: 512 pixels<br>Input number of channels: 3 (RGB values) |
| Below threshold | – | Tumor |
| Above threshold | Region | – |
| Channel (default) | Red | Red |
| Filter (default) | Gaussian (sigmaX: 8.0, sigmaY: 8.0) | Gaussian (sigmaX: 8.0, sigmaY: 8.0) |
| Threshold | 80<br>(pixel intensity value > 80 are classified as region) | Custom threshold |
| minArea | 20,000 $\mu m^2$<br>(minimum area of regions to retain) | 10,000 $\mu m^2$<br>(minimum area of regions to retain) |
| minHoleArea | 8,000 $\mu m^2$<br>(minimum area of connected 'hole' regions to retain) | 8,000 $\mu m^2$<br>(minimum area of connected 'hole' regions to retain) |
| options | "DELETE_EXISTING"<br>(delete existing annotations)<br>"INCLUDE_IGNORED"<br>(include ignored objects) | "DELETE_EXISTING"<br>(delete existing annotations) |

##### Stain Vector Estimation

Next, the stain and background vectors were automatically estimated within the segmented region and updated for subsequent analysis. The new stain vectors were calculated using the EstimateStainVectors function from QuPath. First, the image was smoothed to reduce noise and artifacts. Then, the most frequent RGB values were extracted to determine the background color. Finally, the stain vectors were estimated from the smoothed image, computing the new hematoxylin, DAB, and residual stain components.

##### Threshold-Based Tissue Segmentation

In this step, to optimize the preliminary tissue segmentation a custom threshold was set on the red channel based on the new estimated stain vector values. For images with very light stains, which complicate tissue foreground segmentation, a slightly adjusted custom threshold was defined if the red value of the Hematoxylin stain vector is greater than or equal to 0.7. Additionally, these cases were automatically flagged, and image-level normalization was applied instead of tile-level normalization during cell segmentation. This adjustment addressed background segmentation issues that could have caused local normalization errors. A new pixel classifier, using the updated custom threshold, was then applied to segment the tissue in the slide, computing the final tissue segmentation. Similarly to the preliminary tissue segmentation, the minArea and minHoleArea values were determined through trail and error to obtain a balance between reducing noise and preserving tissue regions. The parameters of the threshold-based tissue segmentation are summarized in Table 2.

##### Cell Segmentation and Classification

The StarDist deep learning model was employed for cell segmentation [24, 25, 26]. More specifically, pixel intensity normalization was performed between the 1st and 99th percentiles of each tile, allowing the model to adapt to the characteristics of each tile. The probability threshold for cell detection was set to 0.25, the pixel size was specified to 0.5 *µm*, and cell overlapping is ignored. Lower values did not improve the results, increasing computational demand, and higher values led to less precise segmentation. Furthermore, the nuclei’ shape and intensity measurements were saved in the QuPath project in case visualization was desired.

The cell classification between positive and negative cells was performed by defining an adaptive threshold based on the triangle thresholding method, which considered the color variations in each image. The DAB-stained images were downsampled by a factor of 4 while preserving high resolution and then were processed using the thresholding module, adapted from Yau Lim [27]. Subsequently, an object classifier was employed to categorize cells equal or above the adaptive threshold as positive and those below as negative, with classifier parameters described in Table 3. Once the classifier was applied, negative cells were displayed in blue, whereas positive cells appeared in red, and relevant data were stored for further post-processing and visualization of the cell density maps.

**Table 3.** Cell classifier parameters.

| Parameter | Value |
| --- | --- |
| Object Classifier Type | SimpleClassifier |
| Classification Method | ClassifyByMeasurementFunction |
| Measurement Used | DAB: Mean |
| Threshold | Adaptive |
| (Object) Filter | DETECTIONS_ALL |

##### Saving Additional Raw Data

Positive and negative cell density maps were generated using the classified nuclei with a search radius of 100 pixels 50*µm*, which defines the surrounding area that contributes to the density value at each point. In addition, the annotation area in *mm*^2^ and the total cell detections of each WSI were stored in .txt files for further processing.

#### 2.2.2 Post Processing in Python

In the Python script, data generated from QuPath was loaded and arranged to provide statistics and visualize results. Summary tables and box plots present findings by diagnosis, including positive and negative cell counts, total cell density, and Ki-67 LI, which is defined as:

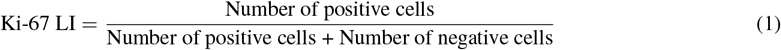

Additionally, the raw grayscale cell density maps generated by QuPath are loaded and processed by normalizing and visualizing them with a colormap, and the Ki-67 LI map is computed as the ratio of positive to total cells to improve interpretation.

### 2.3 Statistical Analysis

The non-parametric Kruskal-Wallis test was conducted to investigate whether a statistically significant correlation existed between the tumor types/families and Ki-67 LI, positive cell density and negative cell density, and tumor descriptors. The significance level was set at *α* = 0.05 and adjusted using the Bonferroni correction.

## 3 Results

All slides were processed in a single round, and the quality of tissue segmentation, cell segmentation and classification was visually assessed after the analysis by at least one non-pathologist, and selected images with artifacts or doubtful tissue structure were revised by a pathologist. Based on the pathologist’s guidance, a total of 26 WSIs were excluded due to artifacts such as background staining, noise on the glass background, light exposure, out-of-focus images, and tissue containing a significant amount of bone fragments, affecting the stain vector estimation, and leading to poor cell classification despite cells being well-segmented. In Figure 3, examples of the excluded cases and a WSI with control tissue are depicted.

### 3.1 Ki-67 WSIs Analysis

In Figure 4, the box plot illustrates the distribution of Ki-67 LI, while Table 4 provides the median, mean ± standard deviation, maximum and minimum values across the tumor families/types. Medulloblastoma exhibits the highest median Ki-67 LI value (19.84), with a mean of 23.10 ± 16.15 and a maximum of 68.75, and ATRT follows with a median of 19.36 and a mean of 20.48 ± 11.2. DIPG, HGG, and ependymoma display moderate Ki-67 LI values, with medians of 11.50, 9.50, and 5.88, respectively. Meningioma had a lower Ki-67 LI, with a median of 1.84 and a mean of 3.37 ± 3.92. The lowest Ki-67 LI values were observed in LGG (median: 0.85), DNET (median: 0.63), and ganglioglioma (median: 0.50). The statistical test suggested a significant correlation between Ki-67 LI and most tumor families/types, with DNET and meningioma not showing a statistically significant correlation.

**Table 4.** Summary of the number of subjects and WSIs included in the results, median (range), mean ± standard deviation of the Ki-67 LI for each tumor family/type. P-values in bold indicate statistically significant differences in Ki-67 LI between tumor families/types, using Kruskal-Wallis test with significance level adjusted using Bonferroni correction *α* = 0.05*/*9 = 0.00556

| Tumor Family/Type | No. of Subjects (Male/Female) | No. of WSIs | Median [Range] | Mean ± Std | p-value |
| --- | --- | --- | --- | --- | --- |
| MB | 80 (45/35) | 85 | 19.84 [1.36, 68.75] | 23.10 ± 16.15 | **<0.001** |
| ATRT | 20 (13/7) | 21 | 19.36 [0.46, 49.35] | 20.48 ± 11.20 | **<0.001** |
| DIPG | 10 (7/3) | 11 | 11.50 [2.40, 31.30] | 12.69 ± 9.19 | **0.005** |
| HGG | 88 (48/40) | 102 | 9.50 [0.10, 56.02] | 12.78 ± 11.93 | **<0.001** |
| EP | 70 (44/26) | 88 | 5.88 [0.04, 64.09] | 10.24 ± 11.79 | **<0.001** |
| MEN | 23 (13/10) | 33 | 1.84 [0.12, 15] | 3.37 ± 3.92 | 0.267 |
| LGG | 257 (140/117) | 291 | 0.85 [0.02, 16.37] | 1.56 ± 2.18 | **<0.001** |
| DNET | 13 (10/3) | 14 | 0.63 [0.12, 53.40] | 4.77 ± 14.04 | 0.038 |
| GG | 50 (28/22) | 63 | 0.50 [0.13, 41.12] | 1.59 ± 5.18 | **0.005** |
| Total | 611 (327/284) | 708 | — | — | — |

**Figure 4.**
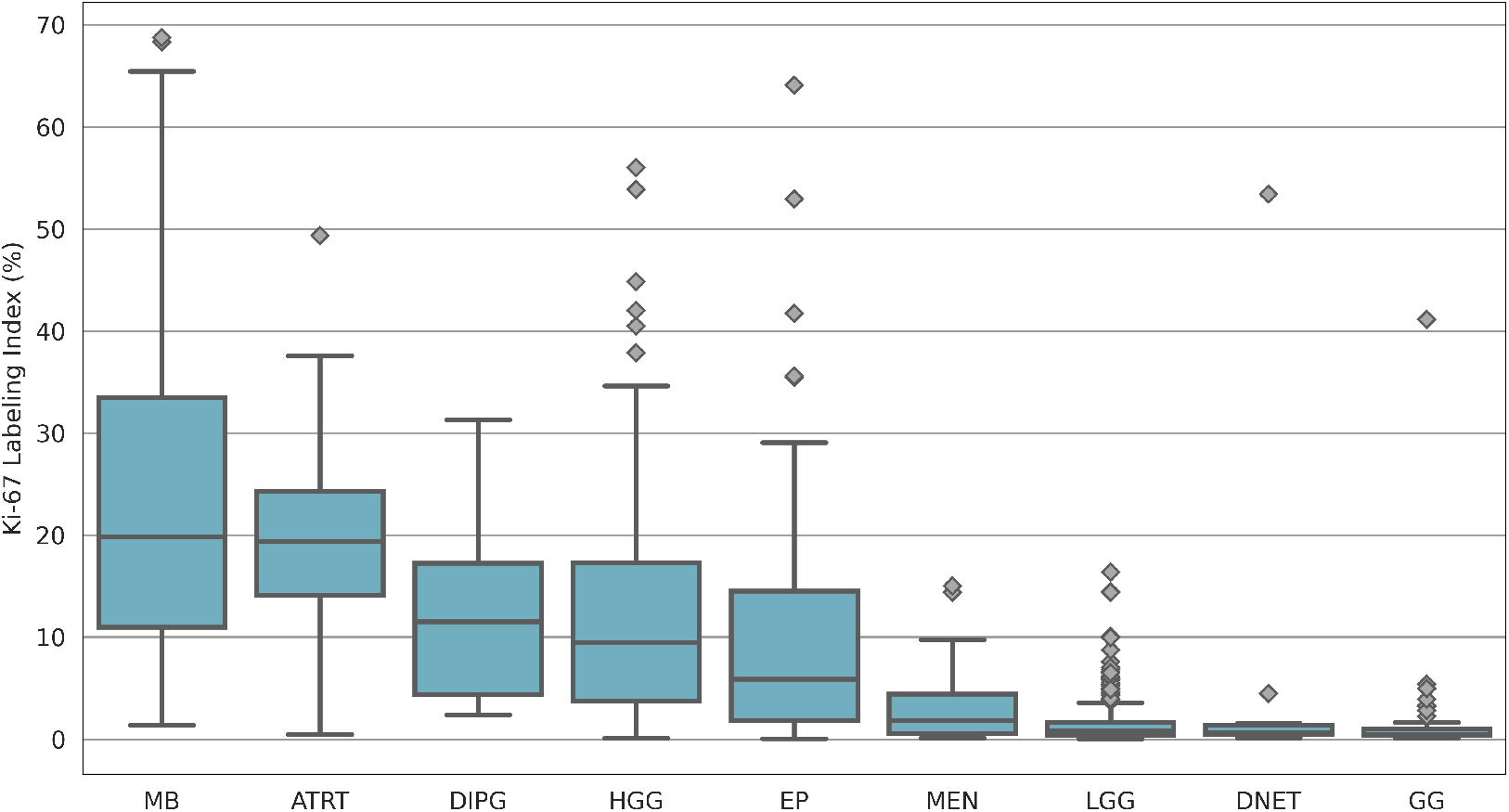
Box plot of Ki-67 LI across the tumor families/types at a WSI level.

Figure 5 presents a logarithmic-scaled box plot illustrating the distribution of positive and negative cell densities (number of positive/negative cells per *mm*^2^) across the tumor families/types, with corresponding statistical values summarized in Table 5. Among the tumors, medulloblastoma exhibited the highest positive and negative cell densities, with medians of 1582.44 cells/*mm*^2^ (mean: 1725.49 ± 1098.42) and 6226.67 cells/*mm*^2^ (mean: 6148.9 ± 2350.98), respectively. ATRT followed closely with a median of 1382.78 positive cells/*mm*^2^ (mean: 1522.89 ± 807.12) and 5908.83 negative cells/*mm*^2^ (mean: 5907.57 ± 1921.75). Ependymoma and HGG demonstrated positive cell density medians of 368.35 and 340.63 cells/*mm*^2^, and negative cell density medians of 5080.68 and 3233.67 cells/*mm*^2^, respectively. DIPG showed slightly lower densities, with a median of 315.46 cells/*mm*^2^ for positive and 2598.08 cells/*mm*^2^ for negative cells. In contrast, meningioma exhibited a relatively low positive cell density (median: 62.49 cells/*mm*^2^), but its negative cell density is comparatively higher (median: 4818.41 cells/*mm*^2^), exceeding those of HGG and DIPG. The lowest positive and negative cell densities were noticed in LGG, DNET, and ganglioglioma. The statistical analysis demonstrated a significant correlation between positive/negative cell density and most of the tumor families/types, except for DNET and DIPG, while no significant correlation was noted between positive cell density and meningioma, and between negative cell density and HGG.

**Table 5.** Summary of the median (range), mean ± standard deviation of the positive and negative cell density (cells per *mm*^2^) for each tumor family/type. P-values shown in bold indicate statistically significant correlation between negative/positive cell density (number of positive/negative cells per mm2) and tumor families/types, using Kruskal-Wallis test with significance level adjusted using Bonferroni correction *α* = 0.05*/*9 = 0.00556.

| Tumor Family/Type | Positive Cell Density |  |  | Negative Cell Density |  |  |
| --- | --- | --- | --- | --- | --- | --- |
| | Median Range | Mean ± Std | p-value | Median Range | Mean ± Std | p-value |
| MB | 1582.44<br>[61.97, 5159.88] | 1725.49 ± 1098.42 | **<0.001** | 6226.67<br>[12995.28, 1119.45] | 6148.90 ± 2350.98 | **<0.001** |
| ATRT | 1382.78<br>[20.02, 2682.21] | 1522.89 ± 807.12 | **<0.001** | 5908.83<br>[9526.94, 2688.03] | 5907.57 ± 1921.75 | **<0.001** |
| EP | 368.35<br>[2.58, 2678.02] | 582.50 ± 649.80 | **<0.001** | 5080.68<br>[9381.84, 1141.03] | 4995.47 ± 1756.61 | **<0.001** |
| HGG | 340.63<br>[2.43, 3088.62] | 515.39 ± 540.29 | **<0.001** | 3233.67<br>[10223.09, 281.82] | 3542.78 ± 1701.45 | 0.705 |
| DIPG | 315.46<br>[44.68, 1183.52] | 373.28 ± 328.61 | 0.051 | 2598.08<br>[3357.26, 1543.41] | 2423.40 ± 596.65 | 0.017 |
| MEN | 62.49<br>[6.52, 1212.32] | 196.53 ± 280.65 | 0.695 | 4818.41<br>[7622.89, 2503.90] | 5044.70 ± 1285.82 | **<0.001** |
| LGG | 21.45<br>[0.47, 577.11] | 41.20 ± 59.05 | **<0.001** | 2674.58<br>[8107.55, 381.12] | 2828.13 ± 1225.46 | **<0.001** |
| DNET | 16.21<br>[1.70, 3907.71] | 302.57 ± 1037.99 | 0.018 | 2431.76<br>[3357.26, 1411.35] | 2453.78 ± 638.27 | 0.009 |
| GG | 13.54<br>[2.14, 1652.21] | 50.93 ± 207.85 | **<0.001** | 2376.80<br>[6868.12, 1048.48] | 2540.76 ± 1205.08 | **<0.001** |

**Figure 5.**
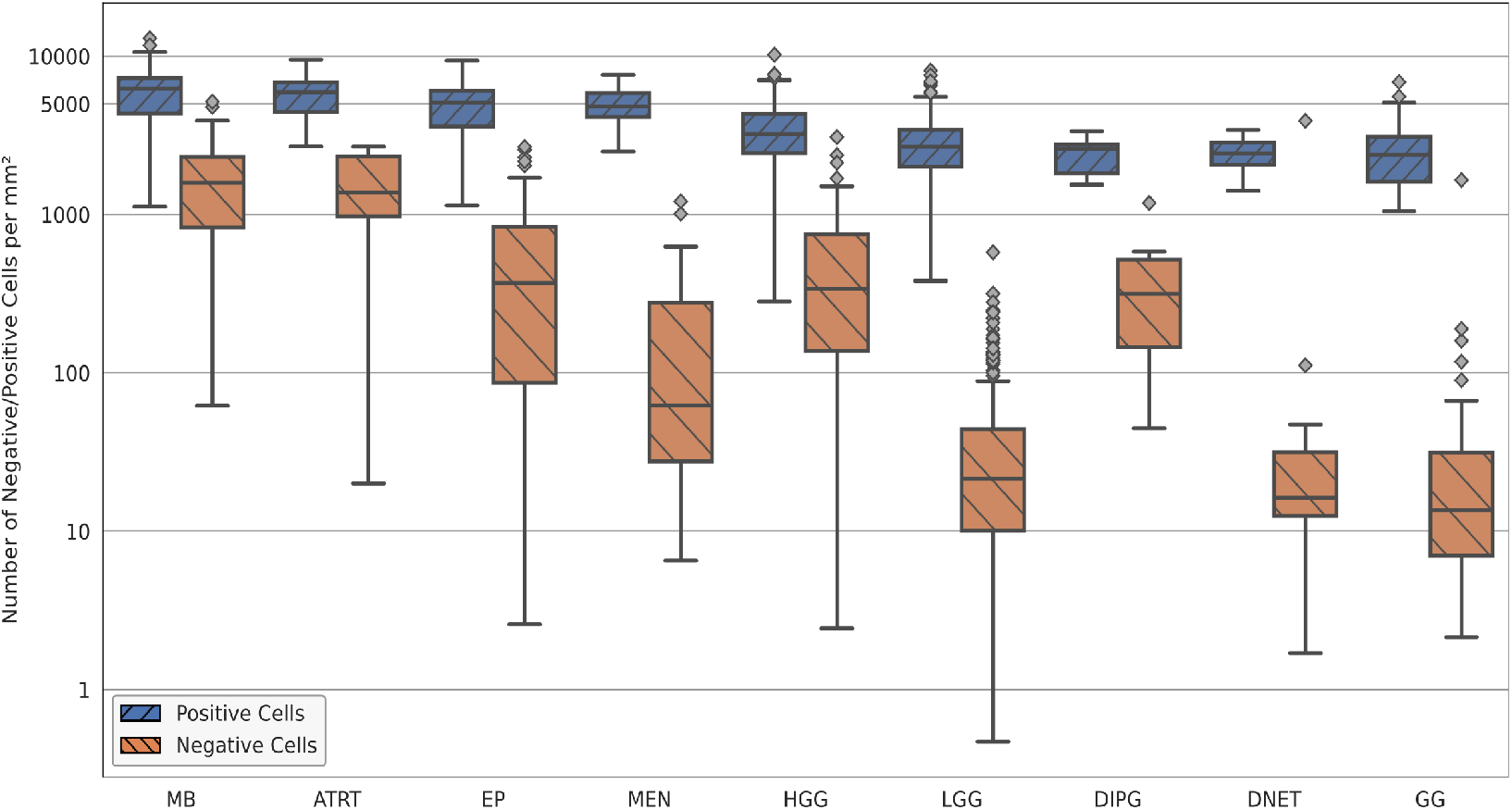
Logarithmic-scaled box plot of the number of the negative and positive cell density (number of negative/positive cells per mm^2^) across the tumor families/types at the WSI level.

Figure 6 describes a logarithmic-scaled box plot of Ki-67 LI across the tumor families/types for each tumor descriptor (after the exclusions, initial CNS tumor: 400, progressive: 85, recurrence: 39, second malignancy: 12). The distribution of Ki-67 LI resembles a similar pattern observed in Figure 4 with no trend being visible within the tumor descriptors. Almost all combinations of tumor families/types and descriptors do not show a statistically significant association with Ki-67 LI. The only notable significant correlations are observed for second malignancy HGG and ependymoma in recurrence. The corresponding p-values are shown in Table 6.

**Table 6.** Results of the Kruskal-Wallis test assessing the statistical correlation between Ki-67 LI and tumor descriptors (initial diagnosis, progression, recurrence, second malignancy) within each tumor family/type at a significance level *α* = 0.05. P-values shown in bold indicate statistically significant correlation between the Ki-67 LI and tumor families/types, using Kruskal-Wallis test with the significance level was adjusted using a Bonferroni correction based on the number of comparisons within each tumor family/type.

| Tumor Family/Type | Tumor Descriptor | No. of Subjects (Male/Female) | No. of WSIs | p-value |
| --- | --- | --- | --- | --- |
| MB | Initial CNS Tumor | 70<br>(41/29) | 70 | 0.927 |
|  | Progressive | 2<br>(1/1) | 4 | 0.667 |
|  | Recurrence | 4<br>(1/3) | 4 | 0.344 |
| ATRT | Initial CNS Tumor | 16<br>(6/10) | 16 | 0.947 |
|  | Progressive | 3<br>(-/3) | 3 | 0.848 |
| HGG | Initial CNS Tumor | 52<br>(24/28) | 54 | 0.680 |
|  | Progressive | 17<br>(8/9) | 21 | 0.232 |
|  | Recurrence | 8<br>(5/3) | 10 | 0.291 |
|  | Second Malignancy | 4<br>(-/4) | 4 | **0.0048** |
| EP | Initial CNS Tumor | 47<br>(30/17) | 54 | 0.346 |
|  | Progressive | 9<br>(5/4) | 10 | 0.989 |
|  | Recurrence | 8<br>(5/3) | 8 | **0.0072** |
| LGG | Initial CNS Tumor | 175<br>(91/84) | 185 | 0.971 |
|  | Progressive | 47<br>(26/21) | 53 | 0.973 |
|  | Recurrence | 16<br>(9/7) | 17 | 0.842 |
|  | Second Malignancy | 3<br>(2/1) | 3 | 0.310 |
| GG | Initial CNS Tumor | 31<br>(16/15) | 41 | 0.919 |
|  | Progressive | 7<br>(5/2) | 7 | 0.877 |
|  | Recurrence | 3<br>(2/1) | 3 | 0.461 |
| MEN | Second Malignancy | 5<br>(1/4) | 5 | 1 |
| DIPG | Initial CNS Tumor | 9<br>(2/7) | 10 | 1 |
| Total | — | 536<br>(280/256) | 582 | — |

**Figure 6.**
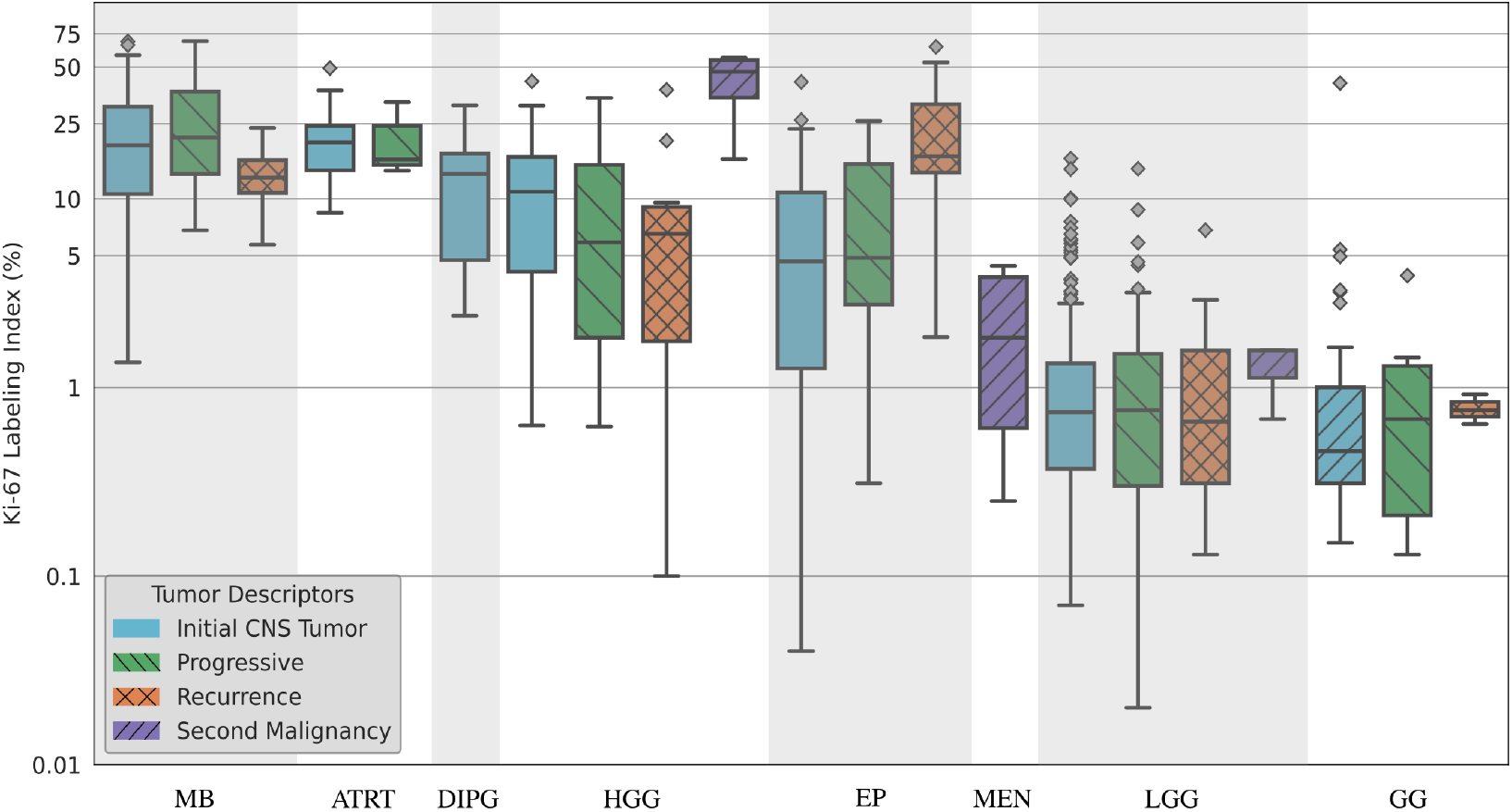
Logarithmic-scaled box plot of Ki-67 LI across the tumor families/types for each tumor descriptor (after the exclusions, initial CNS tumor: 430, progressive: 98, recurrence: 42, second malignancy: 12) at the WSI level. It should be noted that there is no one-to-one correspondence between subjects, diagnosis and tumor descriptors.

### 3.2 Density Maps

The negative and positive cell density maps for each slide were generated by QuPath, which were processed and visualized by the Python script, with the Ki-67 LI map also computed. Figure 7 provides an example of density and Ki-67 LI maps of a subject diagnosed with ATRT with a Ki-67 LI of 49.35. The negative cell density map visualizes the spatial distribution of negatively stained cells in the tissue, with cooler colors (e.g., blue) representing areas with lower negative cell density and warmer colors (e.g., red) indicating regions with higher negative cell density. The color bar associated with this map provides a scale of the density values, helping to interpret the intensity of negative cell density across the tissue. The positive cell density map similarly shows the distribution of positively stained cells, with warmer colors representing higher densities of positive cells. The Ki-67 LI map represents the ratio of positively stained cells to the total number of cells, with warmer colors demonstrating a higher ratio of positive cells and cooler colors showing a lower ratio of positive cells. The color bar provides a scale for the LI, allowing for a clear interpretation of the proportion of positive cells in different tissue regions.

**Figure 7.**
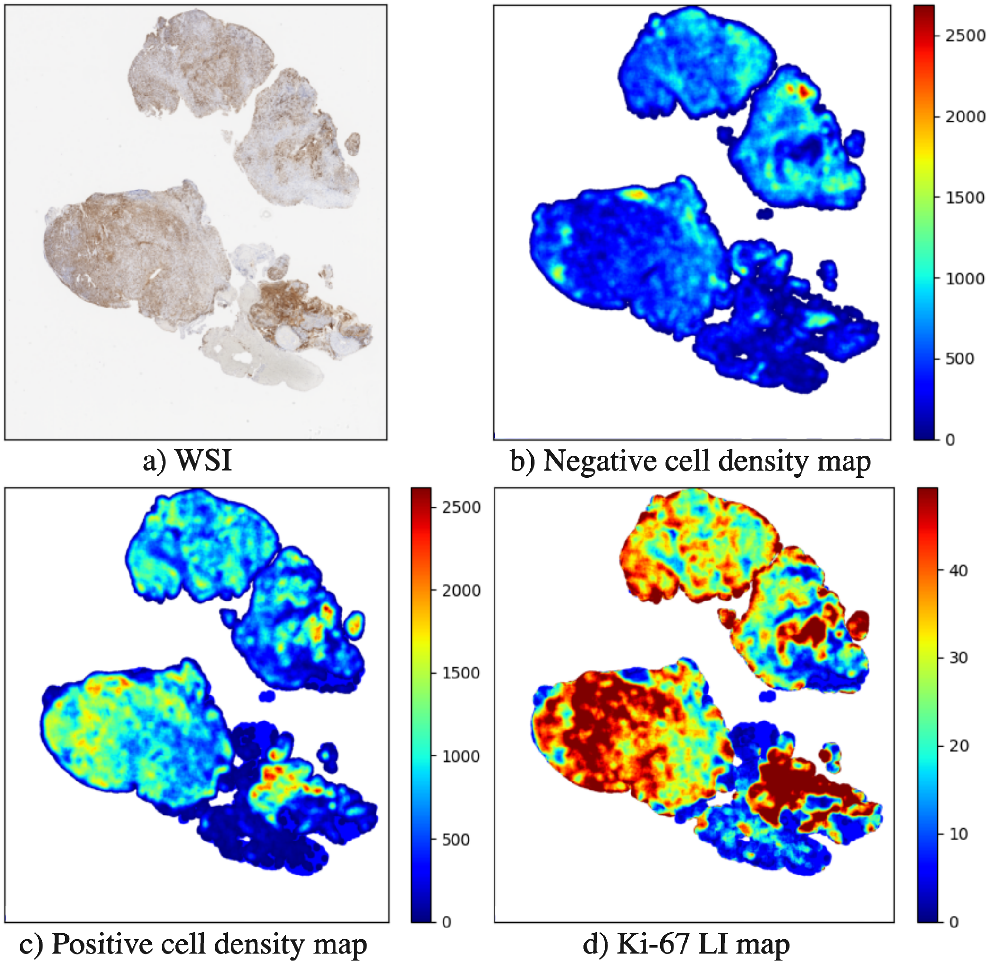
Examples of **a)** WSI for an ATRT tumor with overall Ki-67 LI = 49.35 along with the the corresponding **b)** negative and **c)** positive cell density maps, and **d)** the Ki-67 LI map.

## 4 Discussion

In this study, an Apache Groovy code script for QuPath was developed to automatically calculate the Ki-67 LI on pediatric brain tumor IHC WSIs. Additionally, Python-based algorithms were added to generate summary tables and graphs of the Ki-67 scores and visualize the density maps. The scripts are beneficial for the automatic analysis of relatively large amounts of data using QuPath for research purposes. The codes could be adapted for automated scoring of any other IHC slides for various cancer types, but settings should be adjusted and tested.

The Ki-67 LI values derived from the analysis were aligned with the established oncological consensus, and the statistical analysis confirmed that the Ki-67 LI is a significant marker for distinguishing between different tumor families/types. Specifically, medulloblastomas and ATRTs exhibited considerable higher Ki-67 LI values compared to other tumor families/types, reflecting their higher malignancy. A broad range of Ki-67 LI values is observed in medulloblastomas, possibly reflecting the presence of multiple histological grades. Similarly, the ependymoma cases are related to high Ki-67 LI, although grade 2 and grade 3 tumors might be included, contributing to the variability in the Ki-67 LI. DIPG is typically associated with high Ki-67 LI values, with the definitive diagnosis often requiring molecular testing, as morphological features alone may be insufficient to confirm malignancy. HGG is generally associated with Ki-67 LI values above 10, with some borderline cases being challenging to classify, as low Ki-67 LI values observed in HGG could represent a mixture of low and high-grade areas. The meningioma, LGG, DNET, and ganglioglioma are typically considered low-malignancy tumors, consistent with their correspondingly low Ki-67 LI values. Regarding the relationship between the Ki-67 LI of the tumor families/types and the descriptors, no meaningful insights could be extracted. The absence of a one-to-one correspondence between subjects, tumor family/type, and descriptors limited the feasibility and interpretability of such analysis.

Post-analysis is supported by density maps, allowing pathologists to simultaneously annotate and examine the Ki-67 LI levels in specific regions of interest. The generated Ki-67 LI maps are more practical for highlighting regions with increased positive cell density, simplifying the analysis compared to using only the negative and positive cell density maps. These maps could be used to identify Ki-67 hot spots in WSIs or be compared with explanatory maps (or heatmaps) generated by deep learning models to understand how the model’s attention is localized within tissue regions [28], as demonstrated in this study [29].

Regarding the limitations of the study, the excluded images consisted of cases with severe tissue preparation artifacts, which hinder the accurate visual assessment by a pathologist, as well as cases with visible artifacts, such as bone fragments, which did not significantly impact tissue and cell segmentation but disrupted the stain vector estimation, leading to misleading cell classification. While these cases were relatively few, they still limited the full potential of an automated analysis. Artifacts like pen marks, however, did not interfere with the segmentation and classification of the tissue structure. Furthermore, access to tumor grades for specific types, such as medulloblastoma and ependymoma, would have allowed for a more precise subtype based analysis.

## 5 Conclusion

This study successfully developed and evaluated an automated pipeline for quantifying the Ki-67 LI in pediatric brain tumor WSIs, utilizing an Apache Groovy script in QuPath and a Python-based script for post-processing. The results indicated the effectiveness of this approach in reasonably estimating the Ki-67 LI across various tumor families/types without requiring overly detailed annotations, though a few outlier cases were observed. Additionally, the cell density and Ki-67 LI maps provide a spatial analysis of cell proliferation in pediatric brain tumors, facilitating the identification of areas with high Ki-67 expression or providing automatic annotation for analyzing the explanatory maps of the deep learning models. Overall, these scripts provide a valuable and accessible tool for automatically analyzing large-scale Ki-67 WSI data in research settings, with the potential for adaptation to other cancer types and stains.

## Data Availability

The data can be requested from CBTN (https://cbtn.org).

## 6 Code Availability

The code and detailed instructions for execution linked to this study are available in the GitHub repository: https://github.com/Christoforos-Spyretos/QuPath-Automatic-Cell-Detection-for-Ki-67-WSIs.git.

## 7 Acknowledgements

The research was made possible in part due to The Children’s Brain Tumor Tissue Consortium (CBTTC)/ The Children’s Brain Tumor Network (CBTN). The study was financed by Swedish Childhood Cancer Foundation (MT2021-0011, MT2022-0013), Joanna Cocozza’s Foundation (2025-2026), Linköping University’s Cancer Strength Area (2024), Medical Research Council of Southeast Sweden (FORSS-1011571).

